# Integrative Genomic Analysis Identifies the Soluble Receptor for Advanced Glycation End Products as Putatively Causal for Rheumatoid Arthritis

**DOI:** 10.1101/2020.07.01.20144352

**Authors:** Gha Young Lee, Chen Yao, Shih Jen Hwang, Roby Joehanes, Dong Heon Lee, R. Curtis Ellison, Lynn L. Moore, Chunyu Liu, Daniel Levy

**Author notes:** **Correspondence:** Daniel Levy, MD, Framingham Heart Study, 73 Mt. Wayte Avenue, Framingham, MA 01702.

## Abstract

**Objectives:** Identifying causal biomarkers of rheumatoid arthritis (RA) to improve treatment and monitor disease progression remains a critical but elusive goal. To search for putatively causal protein biomarkers of RA, we designed an integrative genomic strategy utilizing Mendelian randomization (MR), which allows for causal inference between an exposure and an outcome by incorporating genetic variants associated with an exposure (circulating protein level) and inferring its effect on the outcome (rheumatoid arthritis).

**Methods:** We utilized genetic variants associated with 71 cardiovascular disease-related proteins measured in nearly 7000 Framingham Heart Study participants in conjunction with variants associated with RA in a genome-wide association study (GWAS) from the UK Medical Research Council Integrative Epidemiology Unit (19,234 cases, 61,565 controls) to identify putatively causal proteins for RA. In addition, we applied MR to study circulating rheumatoid factor (RF) levels using GWAS of RF from the UK Biobank (n=30,565) as the outcome.

**Results:** We identified the soluble receptor for advanced glycation end products (sRAGE), a critical inflammatory pathway protein, as putatively causal and protective for both RA (odds ratio per 1 standard deviation increment in inverse-rank normalized sRAGE level=0.482; 95% confidence interval 0.374-0.622; p=1.85×10^−08^) and RF levels (β [change in RF level per sRAGE increment]=-1.280; SE=0.434; p=0.003).

**Conclusions:** By integrating GWAS of 71 cardiovascular disease-related proteins, RA, and RF, we identified sRAGE as a putatively causal protein protective for both RA ad RF levels. These results highlight the *AGER*/RAGE axis as a promising new target for RA treatment.

## Introduction

Rheumatoid arthritis (RA) is one of the most common chronic autoimmune diseases with a worldwide prevalence of 0.5% to 1% in adults[1]. Risk factors for RA include a strong genetic component[2], prompting large-scale genome-wide association studies (GWAS) that have revealed more than 100 RA-associated genetic loci[2].

RA is a risk factor for cardiovascular disease (CVD) and multiple studies demonstrate a 1.5-2 fold risk of coronary artery disease in RA patients[3]. Currently hypothesized mechanisms for the predisposition to CVD among RA patients include shared genetic and environmental risk factors[4] and dysregulation of inflammation and immune function[5]. Given their shared features, we investigated a panel of CVD-related plasma proteins for causal effects on RA.

To this end, we analyzed causal relations of 71 CVD-related proteins to RA using protein quantitative trait loci (pQTLs; genetic variants associated with protein levels) from GWAS of plasma protein levels in 6,861 Framingham Heart Study (FHS) participants[6] in conjunction with large-scale GWAS of RA[7] and circulating rheumatoid factor (RF) levels[8], which reflects anti-IgG immunoglobulins present in about 80-90% of RA patients[9].

Mendelian randomization[10] (MR) is an analytical approach to infer causality of an exposure to an outcome by mimicking randomized control trials using genetic variants as instrumental variables (IVs). We applied two-sample MR[10] to identify proteins causally associated with RA and RF. We hypothesized *a priori* that this integrative genomics approach could highlight promising targets for the treatment of RA.

## Methods

### Study Design

The study consisted of three steps (Figure 1). First, from over 16,000 pQTLs identified from GWAS of 71 CVD-related proteins measured in 6,861 FHS participants[6], we characterized pQTL variants that coincided with genetic variants from GWAS of RA[7, 10]. Second, using *cis-*pQTL variants (i.e. residing within 1 Mb of the protein-coding gene) as IVs, we conducted MR testing to infer causal effects of proteins on RA (Figure S1). Third, any causal protein from RA MR analysis was subject to MR analysis investigating its causal effect on RF levels.

**Figure 1.**
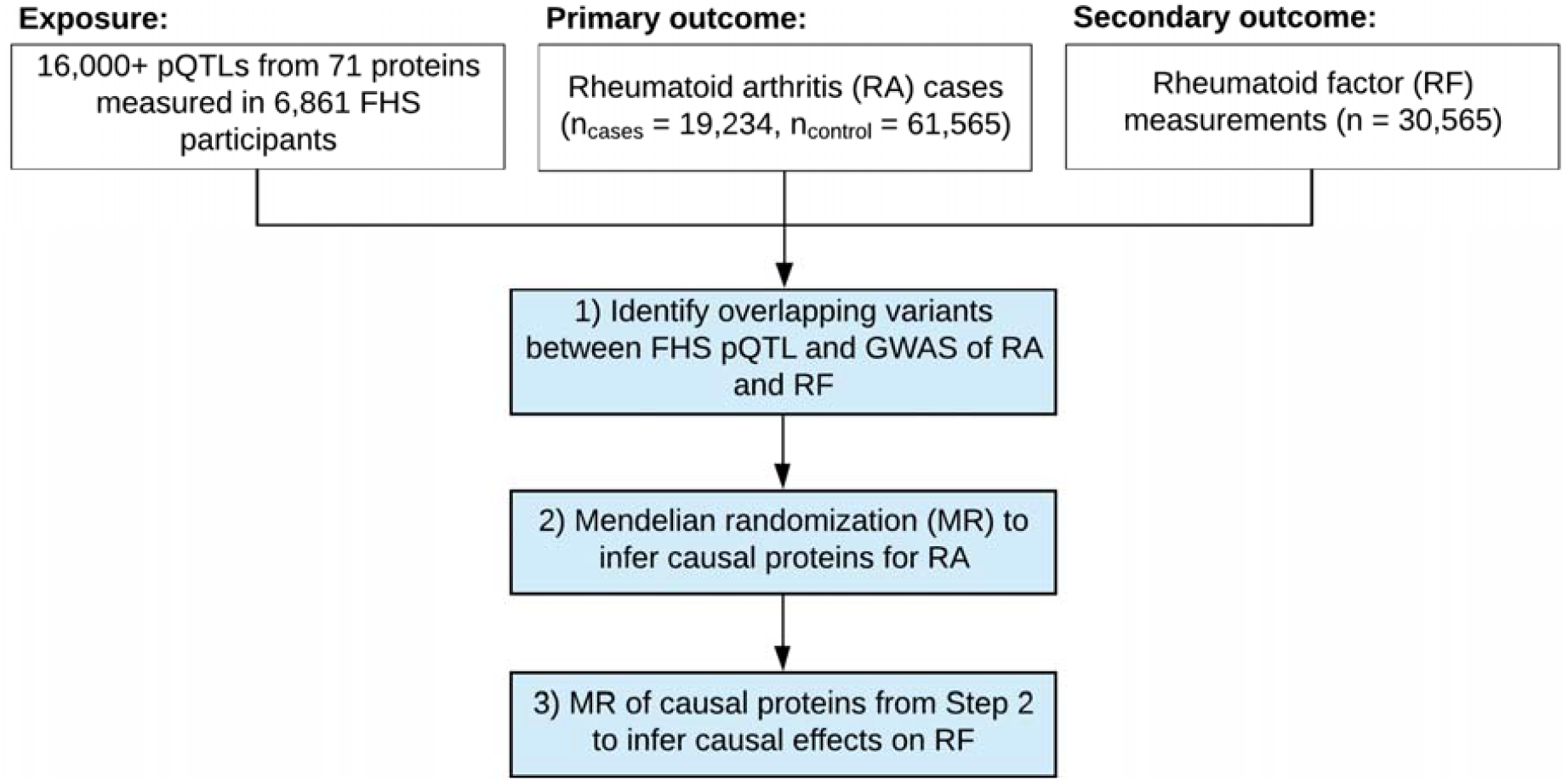
Study Design. Flowchart of the study design. The study consisted of three steps – i. Identify pQTL variants overlapping with genetic variants for RA from GWAS, ii. and iii. Mendelian randomization analyses of the primary and secondary traits. The GWAS for RA was obtained via MRC-IEU[7, 10] and the GWAS for RF[8] was obtained via the UK Biobank.

**Figure 2.**
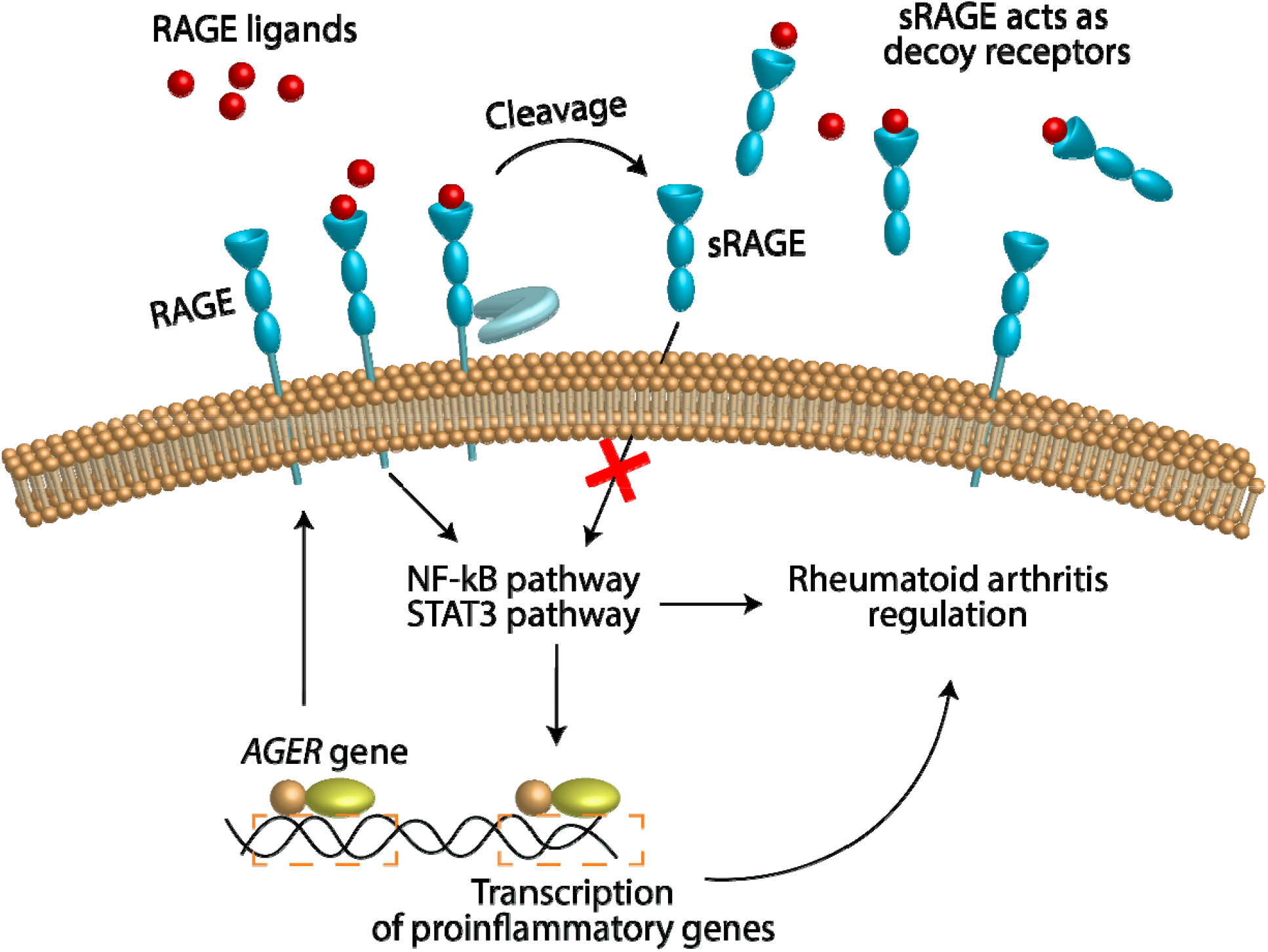
The Protective Role of sRAGE in Relation to Rheumatoid Arthritis. Depiction of the protective mechanism of sRAGE in relation to RA. RAGE is a membrane-bound receptor that triggers pro-inflammatory pathways implicated with RA. sRAGE, a circulating form of RAGE, acts as a decoy receptor for RAGE ligands and therefore downregulates pro-inflammatory pathways.

The 71 plasma proteins were selected based on their presumptive relation to CVD as described previously[6]. Protein levels in FHS participants were measured using Luminex bead-based assays (Luminex, Inc, Austin, TX)[11]. Genotyping of participants was performed using Affymetrix arrays (Affymetrix, Inc, Santa Clara, CA) as well as the Illumina Exome Chip array (Illumina, Inc. San Diego, CA). GWAS of inverse-rank normalized protein levels was performed in R and SAS using genotype dosages based on 1000 Genomes Project imputation (Affymetrix genotypes) or observed genotypes (Exome Chip) in linear mixed-effects models[6].

Our primary analysis used the UK Medical Research Council Integrative Epidemiology Unit’s trans-ethnic GWAS of RA (19,234 cases, 61,565 controls)[7, 10]. All RA cases fulfilled diagnostic criteria of the American College of Rheumatology or were diagnosed by a rheumatologist[7]. Secondary analysis was conducted on the UK Biobank GWAS of circulating RF levels (n=30,565)[8]. The protocols for measuring serum RF and genotyping are described elsewhere[8].

### MR for Causal Inference

Two-sample MR[10] was used to infer causal association between a protein level (using *cis*-pQTL variants as IVs) as the exposure and RA (or RF) as the outcome. Sensitivity analyses were performed to ensure the lack of horizontal pleiotropy[10]. For the exposure, 40 (of 71) proteins with *cis*-pQTLs shared by the outcome GWAS were used[6]. For each outcome, summary statistics were obtained from the corresponding GWAS[7, 8, 10],.

Pruned *cis*-pQTL variants with linkage disequilibrium (LD) of r^2^<0.01 for each protein were used as IVs. For proteins with only one independent SNP after LD pruning, causal effect was determined using the Wald test, *i*.*e*., a ratio of effect per risk allele on RA to effect per risk allele on inverse-rank normalized protein levels. When multiple non-redundant pQTL variants were present, multi-SNP MR was conducted using inverse-variance weighted estimates. All MR analyses were conducted using the TwoSampleMR package in R.

### Patient and Public Partnership

While this is an observational study without patient involvement, the GWAS used involved participant recruitment and outcome measurement as described elsewhere[6, 8, 11].

## Results

MR results are summarized in Table 1 for two proteins (p<0.05) and Table S2 in full. Six out of 40 proteins that had overlapping *cis*-pQTLs were removed by the TwoSampleMR algorithm as the pQTL variants contained ambiguous palindromes or null values. Statistical significance was therefore defined as p<0.0015 (0.05/34). sRAGE was causally implicated (odds ratio [OR] per 1 standard deviation [SD] increment in inverse rank-normalized sRAGE levels=0.482; 95% confidence interval [CI] 0.374-0.622; p=1.85×10^−08^) with a protective effect (OR<1) on RA with the sentinel *cis*-variant rs2070600 driving the causal relationship (Figure S2). Single-SNP MR using rs2070600 demonstrated a causal effect of sRAGE on RA (Wald test OR=0.474; 95% CI 0.446-0.505; p=5.25×10^−122^). sRAGE also was significant (β, RF level change per sRAGE increment=-1.280; SE=0.434; p=0.003) in MR of RF levels without horizontal pleiotropy (p_pleiotropy_=0.953). rs2070600 drove the RF causal relationship (Figure S3); single-SNP MR using rs2070600 revealed evidence of causality (Wald test β=-1.263; SE=0.477; p=0.008).

**Table 1.** Mendelian Randomization Results for Rheumatoid Arthritis (p<0.05) and the Corresponding Mendelian Randomization for Rheumatoid Factor. In MR of both rheumatoid arthritis and rheumatoid factor, sRAGE was the only protein biomarker that passed the multiple testing corrected significance threshold. sRAGE was casual and protective (OR<1; Beta<0) in relation to RA and RF. The odds ratio (OR) and 95% confidence interval (CI) for RA are expressed in terms of per 1 SD increment in inverse rank-normalized protein level, and betas and standard errors (SE) for RF reflect RF level differences (IU/mL) per 1 SD increment in inverse rank-normalized protein level.

| **Exposure** | **Rheumatoid Arthritis** |  |  |  | **Rheumatoid Factor** |  |  |  |
| --- | --- | --- | --- | --- | --- | --- | --- | --- |
|  | **n<sub>snp</sub>** | **OR** | **95% CI** | **P Value** | **n<sub>snp</sub>** | **Beta** | **SE** | **P Value** |
| **sRAGE** | 2 | 0.482 | [0.374;0.622] | 1.85E-08* | 3 | -1.280 | 0.43 | 0.003** |
| **sICAM1** | 1 | 1.200 | [1.026;1.403] | 0.02 | 1 | 1.235 | 0.96 | 0.198 |

\* denotes Bonferroni-corrected significance at P<0.00147 (0.05/34).
\*\* denotes Bonferroni-corrected significance at P<0.025 (0.05/2) for sRAGE and sICAM1.

The minor T allele for rs2070600 was associated with 20-50% lower sRAGE levels in FHS participants (Table S3). The minor allele was associated with increased risk of RA and higher RF levels in the corresponding GWAS[7, 10] (OR (per risk allele)=1.700; 95% CI 1.626-1.777; p=3.60×10^−127^, and β (RF change per risk allele)=0.899; SE=0.340; p=0.008, respectively).

## Discussion

Using an integrative genomic strategy, we identified sRAGE as a putatively causal and protective protein against both RA and RF. sRAGE is a soluble form of RAGE, a transmembrane protein coded by the *AGER* gene. *AGER* is in the major histocompatibility class III locus, near *HLA-DRB1* and the HLA locus, both of which were reported to be associated with RA[2]. Ligands that bind to membrane-bound RAGE, including advanced glycation end products, S100 proteins, and high mobility group box-1 protein (HMGB1), trigger proinflammatory pathways[12]. The circulating sRAGE is derived from proteolytic cleavage of membrane-bound RAGE (mRAGE) or endogenous secretion of an alternatively spliced isoform (esRAGE). sRAGE acts as a decoy receptor and binds to RAGE ligands without inciting RAGE-mediated inflammatory signaling, explaining its protective effect. Indeed, recent studies found sRAGE-overexpressing mesenchymal stem cells (MSCs) had lowered proinflammatory molecule production and increased immunomodulatory molecule expression, and IL-1Ra-knockout mice transplanted with sRAGE-overproducing MSCs demonstrated reduced inflammatory arthritis[13]. Of note, methotrexate, a first-line RA treatment, acts in part by directly binding to the RAGE ligand HMGB1 to inhibit the HMGB1/RAGE pathway[14].

The genetic variant driving the causal effect of sRAGE, rs2070600, is a missense variant in *AGER* exon 3[12] with higher prevalence in RA patients[15]. The amino acid substitution (Gly82Ser) resides at the ligand-binding domain and increases the affinity for RAGE ligands[12], enhancing proinflammatory signaling. This polymorphism is thought to simultaneously make RAGE less susceptible to cell surface RAGE cleavage[12], reducing the generation of sRAGE. This has the dual effects of increasing RAGE ligand-binding and decreasing availability of sRAGE acting as a decoy receptor for proinflammatory ligands. Consistent with these joint effects, we found that the rs2070600 Ser (versus Gly) substitution was associated with lower circulating sRAGE levels in Framingham participants[6] and positively associated with both RA and RF levels in the corresponding GWAS[7, 10].

Previous MR studies of RA have reported IL-6[16] and CRP[17] as causal biomarkers of RA. While IL-6 was not in our panel of proteins, we found that CRP was not causal for RA (p=0.512; Table S2).

Our study has several limitations. First, while we utilized the FHS pQTL variants identified from 71 CVD-related proteins, they are not representative of the entire human plasma proteome. Second, the proteins were measured in plasma, which may yield conclusions not translatable to tissue-specific protein effects. While circulating sRAGE levels are correlated with synovial fluid sRAGE levels (r_s_=0.48, p=0.0002)[18], our findings should be confirmed in tissue-specific settings. Third, while MR testing allowed inference of causal effects of protein levels on RA and RF, further cell and animal studies are warranted. If our findings are confirmed, it may be worthwhile to evaluate the clinical utility of *AGER/*RAGE modulation as a means of reducing inflammatory signaling – e.g. altering cleavage of mRAGE protein to generate more sRAGE in relation to mRAGE or modifying AGER splicing to generate more esRAGE in relation to mRAGE – as therapeutic interventions.

### Key Messages

- Using pQTLs as instrumental variables for 71 CVD-related proteins in conjunction with GWAS of RA and RF, sRAGE was identified as a putatively protective protein.
- sRAGE was previously identified as a potential inhibitor of RAGE-mediated inflammation related to RA.
- Based on our findings, we hypothesize that modulating the *AGER*/RAGE axis is a promising target for RA drug development.

## Data Availability

The pQTL and GWAS used in this study are publicly available from the supplementary files of https://www.nature.com/articles/s41467-018-05512-x and from https://gwas.mrcieu.ac.uk/datasets/, respectively. The results of this study are included in the attached supplementary file in full.

https://gwas.mrcieu.ac.uk/datasets/

## Acknowledgments

The Framingham Heart Study laboratory work for this project was funded by National Institutes of Health contract N01-HC-25195. The analytical component of this project was funded by the Division of Intramural Research, National Heart, Lung, and Blood Institute, National Institutes of Health, Bethesda, MD (D. Levy, Principal Investigator).

## Disclaimer

The views expressed in this manuscript are those of the authors and do not necessarily represent the views of the National Heart, Lung, and Blood Institute; the National Institutes of Health; or the U.S. Department of Health and Human Services.

